# The Experiences of Patients with Vanishing Twin Syndrome: A Mixed-Methods Exploration of Patient Satisfaction and Miscarriage Information

**DOI:** 10.1101/2024.10.04.24314916

**Authors:** Nichole M. Cubbage, Carly Levy, Nicholas Embleton

## Abstract

**Background:** Vanishing Twin Syndrome is a phenomenon where one or more foetuses appear to ‘vanish’ during pregnancy, although in two out of three cases of Vanishing Twin Syndrome there is not complete vanishment of a foetus. Moreover, despite recognition of the syndrome since 1945, a lack of comprehensive guidelines and protocols for its management remains, which may create challenges in patient care and support.

**Objectives:** To explore the experiences of individuals diagnosed with Vanishing Twin Syndrome and analyse common themes among diagnoses and patient-provider communication regarding potential risks and symptoms during pregnancy.

**Study Design:** A global, online survey was created with Qualtrics and comprised 11 questions, including quantitative demographic questions, open-ended qualitative questions, and a sliding-scale rating question. Participants eligible for the study were individuals over the age of 18 who were previously diagnosed with Vanishing Twin Syndrome, currently experiencing Vanishing Twin Syndrome, or believed they may have experienced Vanishing Twin Syndrome without a formal diagnosis. Data analysis utilized Qualtrics’ Stats iQ and Crosstabs iQ tools.

**Results:** Participants reported negative sentiments regarding provider communication of Vanishing Twin Syndrome risks and symptoms, with an average sentiment score of -0.7. Over 53% of responses ranked as -1 on the sentiment scale. The average rating of general information received during Vanishing Twin Syndrome diagnosis was 3.5 out of 10. More than 43% of respondents were not informed of the chorionicity during their Vanishing Twin Syndrome pregnancy despite the known potential impacts of chorionicity on the surviving foetus(es). Discrepancies in information provision were observed across multiple countries, with differences resulting from variations in the quality of patient education and support.

**Conclusion:** The study highlights challenges in communication and support for individuals diagnosed with Vanishing Twin Syndrome and emphasizes a need for improved patient education and guidelines for optimal care. Addressing gaps in patient-provider communication and support may lead to better outcomes and experiences for patients diagnosed with Vanishing Twin Syndrome, as well as their families. Further research is warranted to explore long-term implications and develop tailored interventions for the management of Vanishing Twin Syndrome pregnancies that are also beneficial for patients who experience other forms of miscarriage and infant death in both singleton and multiple pregnancies.

**Tweetable statement:** Study reveals challenges in patient-provider communication and support for individuals diagnosed with Vanishing Twin Syndrome, highlighting a need for enhanced protocols and guidelines for optimal care.

**A. Why was this study conducted?:**

- To explore the experiences of individuals diagnosed with Vanishing Twin Syndrome and analyse common themes
- To assess communication between healthcare providers and Vanishing Twin Syndrome patients regarding potential risks, symptoms, and diagnosis.

**B. What are the key findings?:**

- Negative sentiments of provider communication of Vanishing Twin Syndrome risks and symptoms were commonly reported.
- Low average rating of information received during Vanishing Twin Syndrome diagnosis.
- Patient information provision varied significantly between countries.

**C. What does this study add to what is already known?:**

- Challenges in patient-provider communication and support for individuals diagnosed with Vanishing Twin Syndrome are common.
- Improved support during Vanishing Twin Syndrome diagnosis and pregnancy may improve patient satisfaction
- Identifies gaps in existing guidelines and protocols for the management of Vanishing Twin Syndrome pregnancies.

## Introduction

Vanishing Twin Syndrome (VTS) occurs when one or more foetuses ‘vanish’ from the womb, leading to various outcomes depending on factors such as foetal death cause and gestation period. The three main outcomes of VTS observed include: resorption of deceased foetuses by surviving ones (hence ‘vanishing’), the development of a blighted ovum (i.e., empty sac), and the calcification and compression of deceased foetuses against surviving ones (i.e., *foetus papyraceous*) (Zamani & Parekh, 2021). Despite the three potential outcomes of vanishing twin syndrome (VTS), none guarantees complete disappearance of the deceased foetus(es) (Zamani & Parekh, 2021). VTS can occur in any trimester but is most common in the first (Zamani & Parekh, 2021). However, if VTS occurs later, risks to the surviving multiple(s) and the mother are significantly higher than in the first trimester (Sun et al., 2017). Recent research indicates that reabsorption poses developmental and health risks for survivors and mothers, even when foetal deaths occur as early as 6-8 weeks gestation (Davies et al., 2016). These risks are not yet widely recognized on a clinical level (Song et al., 2020; Weitzner et al., 2023).

Existing standards of care reflect the need for improved clinical protocols regarding miscarriage during pregnancy (Weitzner et al., 2023). Research also suggests that communication informing VTS patients, in particular about miscarriage, may not be clear among healthcare providers due to communication skills and sub-/conscious opinions and biases that may impact their emotional views and logical reasoning (Brann et al., 2020; Hayton, 2010). Additionally, what constitutes human remains can vary. This can further exacerbate challenges for ensuring adequate patient information and foetal remain disposal/memorialization.

Gaps in VTS research contribute to policy inconsistencies (Weitzner et al., 2023) Aside from the biologically complex aspects of this type of miscarriage, confusion is further compounded with ambiguity surrounding the very terminology used to denote and discuss the syndrome. For example, while ‘Vanishing Twin Syndrome’ is the most commonly used, it implies relevance only to twins, leading to misunderstandings in the literature (Batsry & Yinon, 2022). Other terms used include ‘Vanishing Twins Syndrome’ (plural) (Batsry & Yinon, 2022; Roberts & Toth, 2020; Sun et al., 2017). Moreover, although the syndrome may appear to be exclusive to twins, VTS occurs in approximately half of triplet pregnancies, over 35% of twin pregnancies, and at least 20% of assisted reproductive techniques (ART) pregnancies (Zamani & Parekh, 2021). This complexity reflects a need for clarity in terminology and information surrounding VTS.

Establishing guidelines that serve pregnancies where there is a loss of one or more foetuses can be challenging as they necessitate the involvement of professionals that comprise the complex system of care coordination (Weitzner et al., 2023). For example, guidelines may include patient options or resources for foetal remains disposal, memorialization, bereavement, and counselling (Cubbage, 2023). However, establishing guidelines can be challenging as the laws and protocols for discerning and handling human remains and the classification of foetal remains as ‘medical waste’ may vary between care providers, institutions, states, and nations (Middlemiss, 2021; Nahidi et al., 2021).

The National Institute for Health and Care Excellence (NICE) in the UK provides evidence-based guidance for health and social care. NICE offers comprehensive maternal care guidelines, covering antenatal, intrapartum, and postnatal care to ensure safe maternity services. While these guidelines do not mention VTS specifically, they address multiple pregnancies in general. E.g., foetal growth monitoring, screening for specific complications (e.g., Twin-to-Twin Transfusion Syndrome), and providing emotional support for expectant parents (Weitzner et al., 2023). Moreover, although the academic literature denotes risks for VTS patients as early as 6-8 weeks gestation, NICE guidelines currently suggest recording chorionicity and amnionicity between 11w2d and 14w1d (National Institute for Health and Care Excellence, 2013). One study found that implementing NICE guidelines reduced twin stillbirths (Khalil et al., 2020). Canada is the only nation to mention Vanishing Twin explicitly in health guidelines. Clinical resources for providers surrounding VTS are scarce. This is further reflected by searching for the syndrome in clinical decision support tools like BMJ’s Best Practice.

As far as the research team is aware, this study is the first to explore the experiences of individuals who experienced Vanishing Twin Syndrome (VTS) by analysing common themes among diagnoses, prognoses, and patient-provider experience. This information will then be used to make suggestions for potential ways to improve patient care and communication. This is important due to aforementioned trends surrounding VTS and patient experiences.

## Materials and Methods

A global, online survey was created and analysed with Qualtrics. The survey contained 11 questions consisting of several quantitative demographic questions, several open-ended qualitative questions, and one sliding-scale rating question. All questions were optional.

The survey was open to participants for three months. During that period, a combination of snowball sampling, voluntary response sampling, and convenience sampling was deployed due to the small population of VTS participants. The researchers distributed the survey in various VTS and twinless twin parent and survivor support groups, as well as via numerous bereavement and research organizations and social media platforms. Organizations included Multiples of America, the International Council for Multiple Birth Organisations, Return to Zero H.O.P.E., the Pregnancy Loss and Infant Death Alliance, and Twins Trust. Many organizations shared the survey publicly (i.e., via their websites and social media platforms) in addition to sharing privately with their members. Social media platforms used included LinkedIn, Facebook, Instagram, X.

The data analysis phase utilized Qualtrics’ Stats iQ and Crosstabs iQ tools to determine the experiences and sentiment of participants. Individuals were eligible for this study if they were over the age of 18, were able to complete an online survey in English, and could claim at least one of the following:

1. They were previously diagnosed with Vanishing Twin Syndrome (VTS) prior to the study;
2. They were currently experiencing VTS at the time of the study; or
3. The believed they may have previously experienced VTS without receiving a formal diagnosis

Institutional approval for this study was provided by the Institutional Review Board at the Massachusetts College of Pharmacy and Health Sciences on May 2, 2024 (reference number - IRB-2022-2023-123) prior to study commencement. Study recruitment period began on February 27, 2024 and ended April 27, 2024. All participants provided informed, written consent virtually via the study survey. The study was deemed exempt as no private health information was asked of participants in the study survey.

## Results

As denoted in Figure 1 of the 153 responses received total (N = 153), more than 60% came from the United States. However, responses also came from Europe, Australia and New Zealand, Central America, and the Middle East.

**Fig 1.**
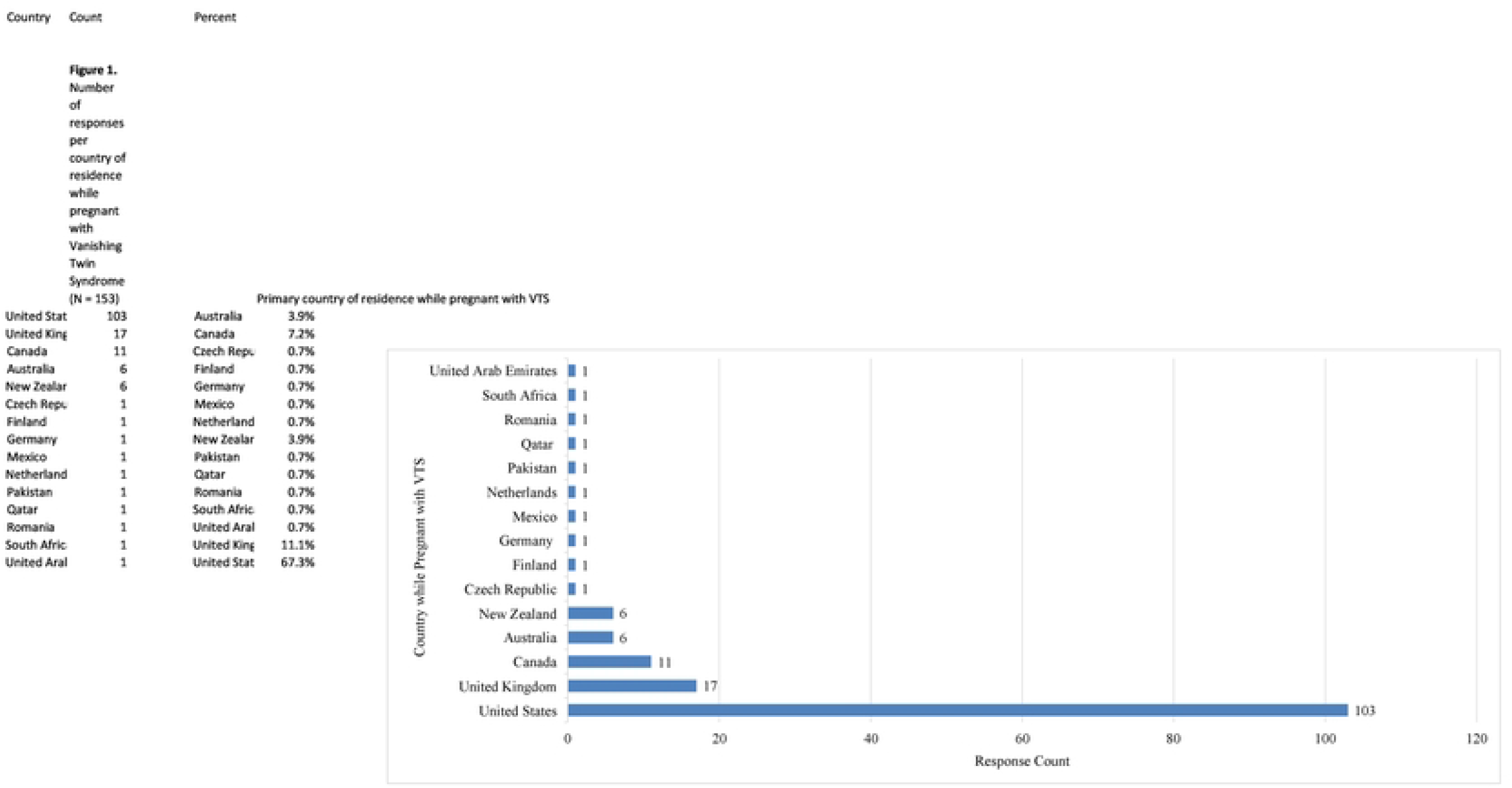

When asked to describe how provider(s) informed participants of any potential risks or symptoms associated with VTS, the average sentiment score was -0.7 with over 53% of responses ranking as a -1 (Table 1). The sentiment ranking scale contains options -2, -1, 0, 1, and 2 (with -2 being the most negative, 0 being neutral, and 2 being the most positive). The standard deviation was 0.8, reflecting a high variation in the quality of responses received from each nation.

**Table 1.** Sentiment analysis of participant recollection of how provider(s) offered information any potential risks or symptoms associated with VTS.

|  |  |
| --- | --- |
| Total Response Count | N = 148 |
| Average Score | -0.7 |
| Median Score | -1.0 |
| Standard Deviation | 0.8 |
| Sentiment Score (-2 = most negative, 2 = most positive) | Percent of responses received per score |
| -2 | 10.1% |
| -1 | 53.4% |
| 0 | 30.4% |
| 1 | 2.7% |
| 2 | 2.7% |

When asked to rate the information received during the VTS diagnosis on a scale of 0-10 (10 being the most informative and 0 being the least). Out of 143 responses from over 15 nations, the average score was 3 with a standard deviation of 2.6, which reflects a high variation in the quality of information received from the patient’s perspective. Countries that scored greater than 4.5 included Romania, South Africa, and the United Arab Emirates with ratings of 6.0, 7.0, and 6.0, respectively. However, each of these countries only had one response. Thus, additional research is needed to determine if the standard deviation is representative of the level of variation that exists between those countries.

When asked if respondents were informed of the chorionicity of their multiples’ during their VTS pregnancy, more than 43% of respondents said ‘no’, as illustrated in Figure 2. Australia had an equal amount of ‘yes’ and ‘no’ responses while just under half of the responses from the United States were marked ‘no’, as noted in Figure 3. Just over 40% of responses from countries comprising the United Kingdom were also marked ‘no’. Responses received from Canada and New Zealand indicate providers in these nations do the best job informing VTS patients of chorionicity based on self-reported patient experiences.

**Fig 2.**
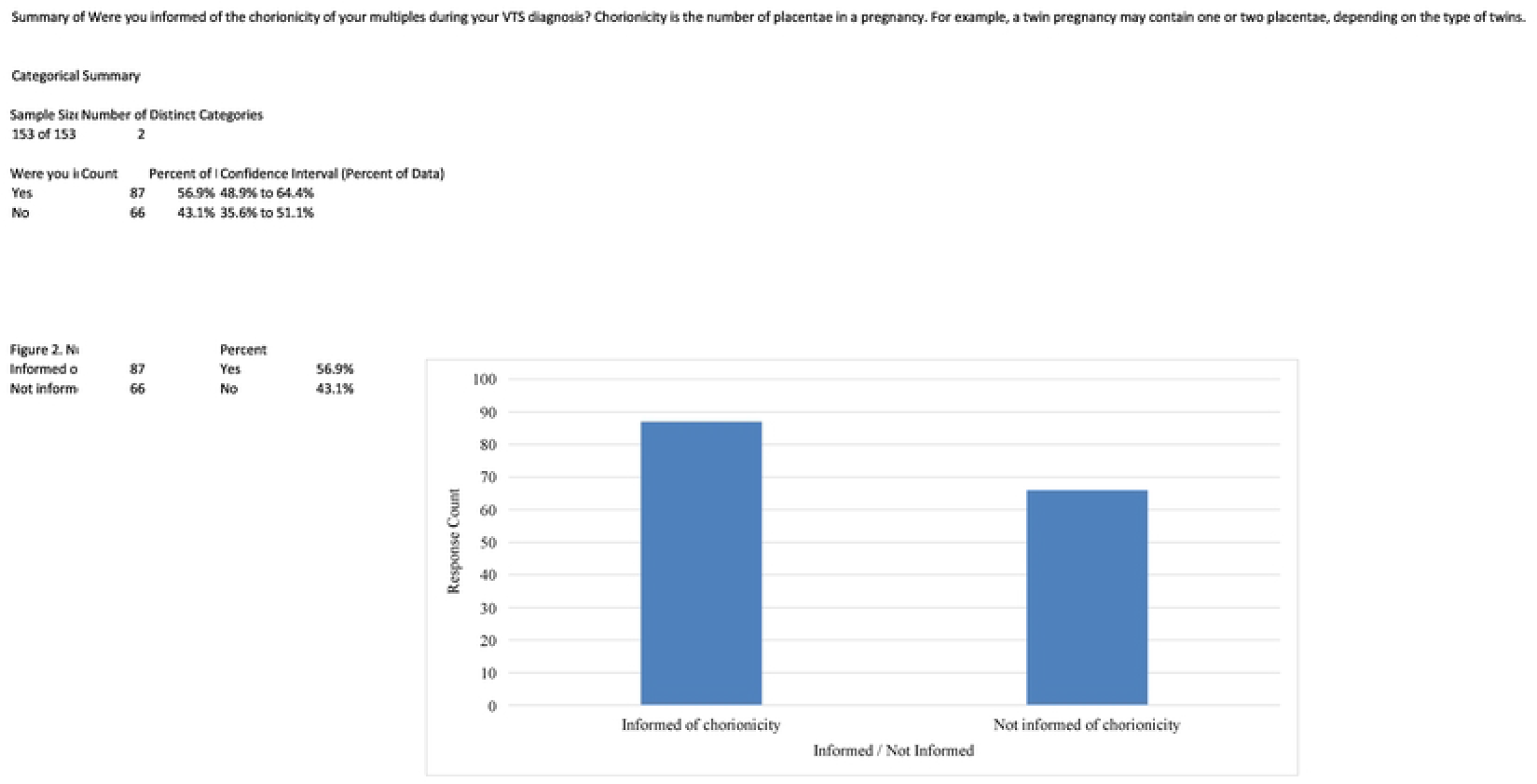

**Fig 3.**
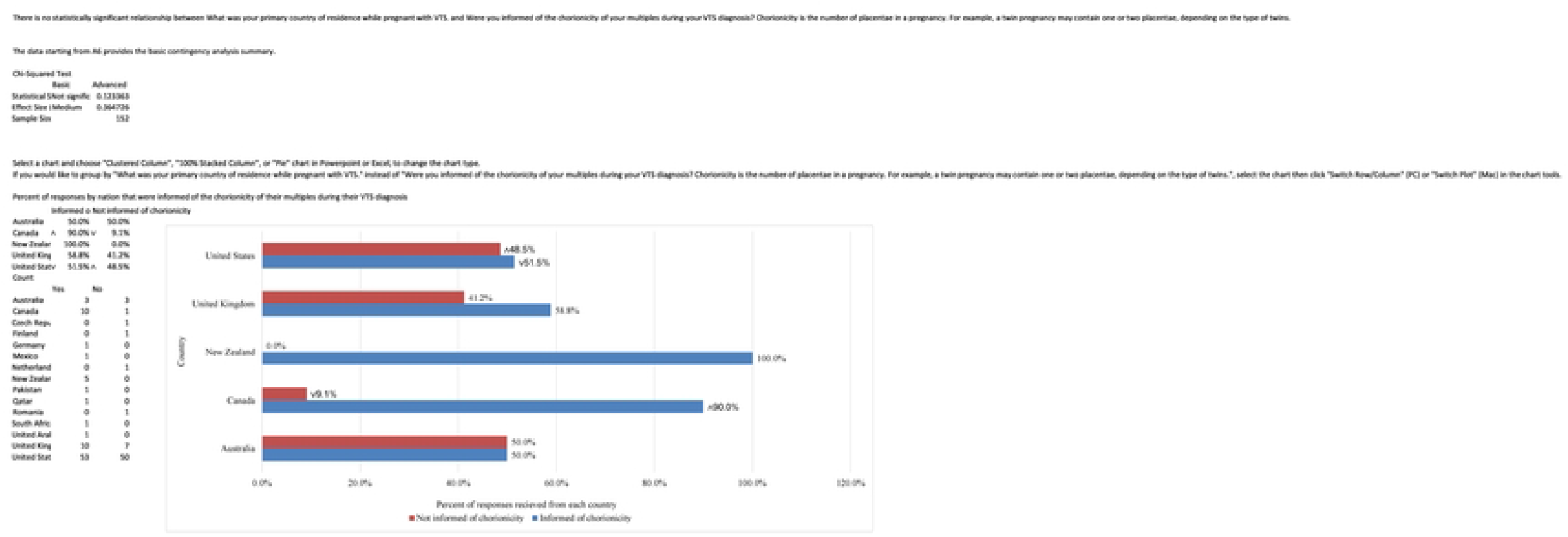

The following five responses are direct quotes gathered from participants in response to the following survey prompt: Describe how your provider(s) informed you of your Vanishing Twin Syndrome diagnosis and the foetal loss(es) that occurred or is currently occurring within your body.

> *They told me at my 7.2 ultrasound that the heart of the second baby was too slow and he was a little behind for his size, so I would be losing him. They booked me another ultrasound 10 days later and his little heart had stopped*.

> *I went in for an ultrasound at 5 weeks pregnant due to bleeding. I was told I was having a di/di twin pregnancy. Both sacs appeared to have implanted “perfectly.” Two weeks later I was on vacation and experienced bright red bleeding. I went to the emergency room and was told there was only one baby and there were no longer any signs of a twin pregnancy. Because it was a different hospital, they did not diagnose me with VTS because they did not have confirmation that my pregnancy had originally been a twin pregnancy. When I returned to my own doctor’s office the next week, I received the VTS diagnosis*.

> *OBGYN told me it was a “piece of junk”. ER Doctor told me I was pregnant with twins but only one had a heartbeat, paperwork showed the chorionicity*.

> *Went to the ER for bleeding early pregnancy thinking I was only pregnant with one baby and miscarrying, ultrasound showed 2 sacs/yolk sacs. I was informed through numerous test results and ultrasound notes*.

> *I was simply shown my living child and the second gestational sack. I had experienced a prior anembryonic pregnancy, prior twin stillbirth, and prior twin live birth, so my provider just acknowledged that the second sac had been a second embryo, but it was not developing*.

Overall, the responses above reflect the diverse ways in which healthcare providers convey the diagnosis of Vanishing Twin Syndrome, from direct communication to instances of confusion due to conflicting information. Additionally, the responses illustrate the complex network of care that can is demanded by obstetrics and gynaecology, especially when there is a reduction in a multiple pregnancy.

Table 2 contains a list of participant responses compiled when asked if there was any information/resource(s) not received during their VTS experiences that they wish they would have.

**Table 2.** Information/resources desired by patients during their Vanishing Twin Syndrome experience.

| **Table 2.** Information/resources desired by patients during their Vanishing Twin Syndrome experience |
| --- |
| Information on support groups |
| Acknowledgement that Vanishing Twin Syndrome is also a miscarriage |
| Acknowledgement of feeling of loss |
| Information on possible long-term effects on surviving child(ren) postpartum |
| Information on risks to surviving fetus(es) in utero |
| Genetic testing information |
| Acknowledgement of physical and mental symptoms |
| Help talking with spouse |
| Grief and loss information/resources |
| Counseling and mental health resources |
| Information on chorionicity/amnionicity |
| Support group information |
| Why Vanishing Twin Syndrome occurred |
| Sex of deceased multiple(s) |
| Information on removal of deceased fetal remains |
| Information on disposal/memorialization/preservation of deceased fetal remains |
| Health risks for mother |
| Better medical imaging of fetus(es) |
| Information on potential implications for future pregnancies |
| Better records/information to refer back to regarding initial diagnosis |
| Better bedside manner from physician to respect loss of child |
| Any information |
| Honesty from providers |

## Comment

### a. Principal Findings

The principal findings of this study reflect challenges faced by individuals diagnosed with Vanishing Twin Syndrome in terms of communication with their respective healthcare providers and the level of information received during diagnosis. Participants reported overall negative sentiments regarding how providers informed them of potential risks or symptoms associated with VTS. Additionally, the study revealed a low average rating of the information received during VTS diagnosis. Together, these novel data indicate a need for improvement in education and support for both provider and patients.

### b. Results in the Context of What is Known

The results presented in this manuscript align with previous literature highlighting the lack of comprehensive guidelines and protocols surrounding VTS diagnosis and management. Despite efforts such as those by the National Institute for Health and Care Excellence in the United Kingdom, explicit mention of VTS in guidelines remains scarce, leading to inconsistencies in patient care and support in healthcare systems across the globe. This study contributes to the existing body of knowledge by being the first to light on VTS patient experiences and highlight areas for improvement in clinical practice.

### c. Clinical Implications

The study findings underscore the importance of enhancing communication between healthcare providers and patients diagnosed with VTS. Improved patient education and support regarding potential risks and symptoms associated with VTS are crucial for ensuring optimal patient outcomes and well-being. Clinicians should strive to provide comprehensive information during the diagnosis of VTS, addressing patients’ concerns and offering appropriate emotional support. Additionally, there is a need for the development of specific guidelines and protocols tailored to the management of VTS pregnancies to ensure consistent and standardized care across healthcare settings.

### d. Research Implications

This study highlights several unanswered questions and areas for future research in the field of VTS. Further investigation is warranted to explore the long-term psychological and physiological implications of VTS on both patients and surviving multiples.

Additionally, prospective studies are needed to evaluate the effectiveness of interventions aimed at improving patient-provider communication and support in the context of VTS diagnosis and management. Longitudinal studies tracking the outcomes of VTS pregnancies and the experiences of individuals diagnosed with VTS may also be able to provide valuable insights into optimal clinical management strategies which will likely vary by jurisdiction and institution.

### e. Strengths and Limitations

Strengths of this study include its global reach and inclusion of participants from diverse geographic regions, providing a broad perspective on patient experiences with VTS. The utilization of both quantitative and qualitative data collection methods allowed for a comprehensive understanding of the topic. However, limitations include the reliance on self-reported data and the sampling methods used, which may be subject to recall bias and sampling bias, respectively. Responses gained throughout this research do not include individuals who do not have access to online surveys. Additionally, the study’s cross-sectional design limits the ability to establish causal relationships between variables. Relationships between variables are also inhibited in some cases as medical education/training, laws, and regulations can vary by nation, state/province, and individual institutions. Moreover, most of the responses gathered were from 1995 – present. Older populations born before routine ultrasound use may have received VTS diagnoses (e.g., postpartum identification of foetus papyraceous) but were less likely. Despite the decline of this population, it is necessary to acknowledge their existence, consider the impacts of technological advancements, and understand how such advancements may impact data availability and results gathered therefrom.

### f. Conclusions

In conclusion, this study highlights the challenges faced by individuals diagnosed with VTS in terms of communication with healthcare providers and the level of information received during diagnosis. Addressing these gaps in clinical practice can lead to better patient-provider communication, and improved patient outcomes and experiences for VTS patients, families, and providers.

## Data Availability

All relevant data are within the manuscript and its Supporting Information files.

## Acknowledgments

1. Multiples of America
2. Dr. Marc A. Nascarella, Employed by: Massachusetts College of Pharmacy and Health Sciences
3. International Council of Multiple Birth Organisations (ICOMBO) and Dr. Carloyn Lister (Chair and Research Director)
4. Dr. Anthony Lacina, Employed by: Massachusetts College of Pharmacy and Health Sciences
5. Dr. Helen Ewing, Employed by: Massachusetts College of Pharmacy and Health Sciences
6. Dr. Kiley Hanish, Founder and President of Return to Zero H.O.P.E.
7. Pregnancy Loss and Infant Death Alliance (PLIDA)
8. Twins Trust
9. Dr. Jeff Craig, Employed by: Murdoch Children’s Institute
10. Dr. Wolfgang Rumpf, Employed by: the University of Maryland Global Campus
11. The Columbus Mothers of Twins Club
12. Alvaro Danilo Silva, Employed by: Silva Music Studios
13. Alison Jacobson, CEO of First Candle
14. Zane Welker, Employed by: Phosphorix Ltd.

None of the above individuals or entities served as funding sources or sources of compensation, whether financial or in the form of services or complimentary products.

During the preparation of this work the author(s) used ChatGPT 3.5 in order to reduce the word count of the original manuscript. After using this tool/service, the author(s) reviewed and edited the content as needed and take(s) full responsibility for the content of the publication.

## Notes

Disclosure of Interest: The authors no conflict of interest.

### Competing Interest Statement

The authors have declared no competing interest.

### Funding Statement

The author(s) received no specific funding for this work.

### Author Declarations

Institutional approval for this study was provided by the Institutional Review Board at the Massachusetts College of Pharmacy and Health Sciences on May 2, 2024 (reference number - IRB-2022-2023-123). The study was deemed exempt as no private health information was asked of participants in the study survey.

